# The persistent underrepresentation of patients with chronic kidney disease in cardiovascular trials: a systematic review and evidence map of exclusion and outcomes

**DOI:** 10.1101/2023.07.18.23292848

**Authors:** Julia M.T. Colombijn, Demy L. Idema, Sanne van Beem, Anna Marthe Blokland, Kim van der Braak, M. Louis Handoko, Linde F. Huis in ’t Veld, Tabea Kaul, Nurda Kolagasigil-Akdemir, Mike P.T. Kusters, Sabine C.A. Meijvis, Ilse J. Oosting, Rene Spijker, Michiel L. Bots, Lotty Hooft, Marianne C. Verhaar, Robin W.M. Vernooij

## Abstract

**Background:** Patients with chronic kidney disease (CKD) are at high risk for cardiovascular disease, but their systematic underrepresentation in cardiovascular randomised controlled trials (RCTs) limits appropriate evidence to guide cardiovascular risk management (CVRM). This systematic review aims to evaluate trends in the underrepresentation of patients with CKD in cardiovascular RCTs in the past 20 years and highlight evidence gaps for CVRM medications in this population.

**Methods:** A systematic search was conducted in ClinicalTrials.gov from its inception in 2000 until October 2021, targeting RCTs evaluating the efficacy of CVRM medications on mortality, cardiovascular disease, and kidney failure in adults with cardiovascular disease or one or more cardiovascular risk factors. Two reviewers independently screened references and extracted data. Outcomes were the exclusion rate of patients with CKD over time and an evidence map of studies reporting results for this population.

**Results:** In total, 1194 RCTs involving 2,207,677 participants were included. Since 2000, the percentage of cardiovascular RCTs that exclude patients with CKD has increased from 66% to 79% (74% overall, 884 RCTs). In 73% of RCTs, more patients were excluded than anticipated on safety grounds (63% without dose adjustment necessary and 79% of RCTs with dose adjustment necessary). In total, 158 RCTs (13%) reported results patients with CKD separately (e.g. in subgroup analyses). Significant evidence gaps exist for most CVRM interventions for patients with CKD, particularly for those with CKD stage 4-5. For patients with an eGFR <30 ml/min/1.73m^2^, 23 RCTs reported results, for dialysis patients 15 RCTs, and for kidney transplant patients only 1 RCT.

**Conclusion:** The underrepresentation of patients with CKD in cardiovascular RCTs has not improved in the past two decades and three-quarters of RCTs excluded more patients than expected on safety grounds. A lack of RCTs that report results for patients with CKD has resulted in significant evidence gaps for most CVRM medications in all subgroups of patients with CKD, in particular for those with CKD stage 4-5.

**Primary funding source:** Dutch Heart Foundation, 2020B008 RECONNEXT

**Registration:** PROSPERO (CRD42022296746)

## Introduction

Chronic kidney disease (CKD) affects almost 700 million patients worldwide and is the cause of 1.9 million cardiovascular deaths annually (1, 2). Over 60% of patients with CKD has a history of cardiovascular disease and this is also the main cause of death in this population (3, 4). Almost all patients with CKD have a much higher risk for CVD than for kidney failure (5, 6). This elevated CVD risk is already observed for patients with an estimated glomerular filtration rate (eGFR) <75 ml/min/1.73m^2^ and increases as CKD progresses, independently of other risk factors like hypertension and diabetes (3, 7).

The high cardiovascular risk in patients with CKD underscores the importance of effective strategies for cardiovascular risk management (CVRM) for patients with CKD. Nevertheless, even though over 90% of patients with CKD are being prescribed CVRM medications, evidence on the safety and efficacy of these medications in patients with CKD is limited (8, 9), Historically, patients with CKD largely have been underrepresented in cardiovascular randomised controlled trials (RCTs). They are frequently excluded due to concerns about the safety and efficacy of interventions. Even the RCTs without explicit CKD exclusion criteria often do not include these patients nor assess treatment effects for them (10–13).

A lack of information about the efficacy of CVRM medications in people with CKD undermines effective CVRM treatment. Efficacy estimates about CVRM medications from RCTs excluding patients with CKD cannot be extrapolated carelessly since the increased CVD risk in patients with CKD and altered pathophysiology of CVD can modify the efficacy of treatments (14). As CKD progresses to kidney failure, patients’ CVD burden shifts from atherosclerotic CVD to medial arterial calcification, cardiac arrhythmias, left-ventricular hypertrophy, and sudden cardiac death (14). A higher cardiovascular risk could enhance the efficacy of CVRM treatments for patients with CKD as a greater absolute risk reduction can be achieved. However, a reduced life expectancy and the induction of additional pathways in CVD pathophysiology which are not inhibited by traditional CVRM medications could offset these benefits and render treatment futile (15).

Several systematic reviews, which included RCTs published up to 2014, have reported on the underrepresentation of patients with CKD in cardiovascular RCTs (10–13). However, it is unclear whether the representation of patients with CKD in cardiovascular RCTs has improved over the past years and whether this population has been included in RCTs evaluating the efficacy of (relatively) new treatments, such as sodium glucose-cotransporter-2 inhibitors (SGLT2-inhibitors) and direct oral anticoagulants (DOACs). Furthermore, the systematic exclusion of patients with CKD makes it uncertain for which CVRM medications evidence is available on the efficacy and safety specifically for people with CKD. An overview of the RCTs evaluating the efficacy of CVRM medications for (different subgroups of) patients with CKD is currently lacking. Therefore, the aim of this systematic review was to evaluate trends in the underrepresentation of patients with CKD from cardiovascular RCTs in the past 20 years, and to create an overview of the available evidence, highlighting evidence gaps for CVRM medications for patients with CKD.

## Methods

This systematic review has been reported according to the Preferred Reporting Items for Systematic Reviews and Meta-Analyses (PRISMA) and registered prospectively in PROSPERO (CRD42022296746) (16). The full protocol has been published previously (17).

### Data sources and searches

In brief, ClinicalTrials.gov was searched through the Cochrane Central Register of Controlled Trials from inception (February 2000) until October 2021 with a combination of keywords for CVD, cardiovascular risk factors and the included interventions to identify planned, ongoing, terminated, and completed RCTs. Next, the full-text publications were retrieved up to May 2023 from ClinicalTrials.gov. If no full texts were listed in ClinicalTrials.gov, MEDLINE, Embase, and Google Scholar were searched additionally to retrieve the full texts. Records of which no publications could be found were excluded. Landmark RCTs not identified in our search were added manually.

### Study selection

Two reviewers screened clinical trial records and publications independently based on the eligibility criteria. Disagreements were resolved by discussion.

Eligible RCTs were those evaluating the effect of antiplatelets, anticoagulants, blood pressure-lowering drugs, glucose-lowering drugs, or cholesterol-lowering drugs recommended by the European Society of Cardiology, the American Heart Association, the American Stroke Association, the American College of Cardiology, and the American Diabetes Association for the prevention of CVD (18–39) on all-cause or cardiovascular mortality, cardiovascular disease (as composite endpoint and individual events), peripheral arterial disease, or kidney failure, in adults with a history of CVD or one or more CVD risk factors. Interventions must be compared against placebo, usual care, another therapy, or a different treatment dosage or duration.

### Data extraction and quality assessment

Data extraction was performed by one reviewer with a standardised extraction form and verified by a second reviewer. A list of extracted variables is described in the protocol (17). Risk of bias was not assessed since bias in study design is unlikely to affect whether patients with CKD are excluded from RCTs or whether authors report results for them (as subgroup analysis or by restriction of the study population).

### Data synthesis and analysis

Outcomes of interest were the frequency of exclusion of patients with CKD and the reporting of results for patients with CKD (through subgroup analyses or restriction of the study population). Exclusion of patients with CKD was defined as the exclusion of patients on kidney-related eligibility criteria. If RCTs did not specify kidney-related eligibility criteria, we presumed these patients were not excluded.

Categorical variables were described as frequency (percentage) and continuous variables as mean (standard deviation) if they followed a normal distribution and otherwise as median [interquartile range]. The frequency of exclusion of patients with CKD was evaluated for different time periods, medications, and dosing recommendations for patients with CKD. Dosing recommendations were categorised based on the Renal Drug Handbook as no dose adjustment necessary, dose adjustment in CKD stage 3, or 4-5, and contra-indicated in CKD stage 4, or 5 (40).

## Results

### Study selection, and characteristics of studies and participants

Overall, 13,017 RCTs were identified in the search of which 8,780 were excluded on ClinicalTrials.gov record. Of 1419 RCTs, no full-text was retrieved. Of the remaining 2818 records screened on full text, 1,194 RCTs involving 2,207,677 participants were included (Supplementary Figure 1). Main reasons for exclusion were no outcomes of interest (n=884), no interventions of interest (n=304), and insufficient sample size (n=77). Included RCTs had a median follow-up of 24 months [IQR 12.0 – 39.6] and 82 (7%) had published a protocol only. Glucose-lowering drugs were evaluated in 552 RCTs (46%), antiplatelets and anticoagulants in 229 RCTs (19%), blood pressure-lowering drugs in 221 RCTs (19%), and a combination of these interventions in 30 RCTs (3%) (Table 1).

**Table 1:** characteristics of included RCTs.

| Variables | All (n=1194) | Exclusion of people with CKD (n=884) | No exclusion of people with CKD (n=310) |
| --- | --- | --- | --- |
| **Study characteristics** |  |  |  |
| Year of publication |  |  |  |
| <2000 | 21(2) | 12 (1) | 9 (3) |
| 2000-2005 | 50 (4) | 33 (4) | 17 (5) |
| 2006-2010 | 239 (20) | 173 (20) | 66 (21) |
| 2011-2015 | 393 (33) | 286 (32) | 107 (34) |
| 2016-2020 | 418 (35) | 322 (36) | 96 (31) |
| >2020 | 73 (6) | 58 (7) | 15 (5) |
| Sample size |  |  |  |
| Total | 2207677 | 1.595.831 | 563.038 |
| Median [IQR] | 568 [304 – 1315] | 559 [311 – 1250] | 530 [280 – 1288] |
| Follow-up (months) | 24.0 [12.0 – 39.6] | 26.4 [12.0 – 40.8] | 17.8 [5.5 – 30.0] |
| Only protocol published | 81 (7) | 64 (8) | 17 (5) |
| Location |  |  |  |
| Multi-continental | 576 (48) | 443 (50) | 133 (43) |
| Europe | 165 (14) | 117 (13) | 48 (16) |
| North America | 172 (14) | 116 (13) | 53 (17) |
| Asia/Australia/Africa/Other | 281 (24) | 208 (24) | 73 (24) |
| Funding source<sup>#</sup> |  |  |  |
| Industry | 987 (83) | 736 (83) | 251 (81) |
| Government | 81 (7) | 57 (8) | 24 (6) |
| Institutional | 151 (13) | 110 (12) | 41 (13) |
| Unspecified/miscellaneous | 49 (4) | 35 (4) | 14 (5) |
| Type of intervention |  |  |  |
| Antihypertensives | 221 (19) | 182 (20) | 39 (13) |
| Glucose lowering drugs | 552 (46) | 424 (48) | 128 (41) |
| Lipid lowering drugs | 162 (14) | 126 (15) | 36 (12) |
| Antiplatelets or anticoagulants | 229 (19) | 132 (15) | 97 (31) |
| Combination of interventions | 30 (3) | 20 (2) | 10 (3) |
| **Participant characteristics** |  |  |  |
| Age (years) | 63 ± 6 | 64 ± 6 | 62 ± 6 |
| Sex (% female) | 36 ± 12 | 37 ± 12 | 33 ± 12 |
| eGFR (ml/min/1.73m<sup>2</sup>)* |  |  |  |
| mean (SD) | 73 ± 13 | 73 ± 13 | 77 ± 10 |
| Not reported | 899 (75) | 623 (71) | 276 (89) |
| ≥90, | 79 (27) | 74 (28) | 5 (14) |
| 60-89 | 169 (57) | 144 (55) | 25 (73) |
| 45-59 | 27 (9) | 24 (9) | 3 (8) |
| 30-44 | 13 (4) | 13 (5) | 0 (0) |
| 15-29 | 7 (2) | 6 (3) | 1 (3) |
| Serum creatinine (mg/dL)* |  |  |  |
| Median [IQR] | 1.00 [0.96 – 1.04] | 1.00 [0.97 – 1.04] | 1.00 [0.95 – 1.01] |
| Not reported | 1040 (87) | 750 (85) | 290 (94) |
| <1.0 | 77 (50) | 67 (50) | 10 (50) |
| 1.0-1.49 | 61 (40) | 51 (38) | 10 (50) |
| 1.5-2.00 | 9 (6) | 9 (7) | 0 (0) |
| >2.0 | 7 (5) | 7 (5) | 0 (0) |
| Dialysis | 17 (<1) | 17 (2) | 0 (0) |
| Transplantation | 1 (<1) | 1 (<1) | 0 (0) |

Abbreviations: CKD, chronic kidney disease; eGFR, estimated glomerular filtration rate; IQR, interquartile range. Categorical variables are describes as n (percentage). Normally distributed continuous variables as mean ± SD and skewedly distributed variables as median [IQR].
<sup>#</sup> RCTs can have multiple funding sources. Therefore percentages do not add up to 100 percent.
\*percentages only for proportion without missing eGFR or serum creatinine.

Participants had a mean age of 63±6 years and the mean percentage of women was 36±12. The mean eGFR was 73±13 ml/min/1.73m^2^ and the median serum creatinine level was 1.00 [0.96-1.04] mg/dl but these variables were reported in only 295 (25%) and 154 (13%) RCTs, respectively. Patients receiving dialysis were included in 17 RCTs (1%) and patients with a kidney transplant in 1 (<1%) RCT. An overview of included RCTs and their characteristics is listed in Supplemental Table 1.

### Trends in the underrepresentation of patients with CKD

Since 2000, the percentage of RCTs that excluded (subgroups of) patients with CKD has increased from 66 to 79% (74% overall, 884 RCTs; Figure 1a). Patients with CKD stage 1-3 (i.e., eGFR >30 ml/min/1.73m^2^, serum creatinine <2 mg/dL or a history of CKD) were excluded in 458 RCTs (38% of all included RCTs and 52% of RCTs which excluded patients with CKD) (Figure 1a). In the past 20 years, patients with CKD stage 4-5 have been excluded from cardiovascular RCTs more frequently while the exclusion of patients with CKD stage 1-3 has remained stable (Figure 1a). The proportion of RCTs where dose adjustment based on kidney function was required or where medication was contra-indicated on kidney function remained consistent across this different time period (Figure 2b, Supplemental Figure 2). The kidney exclusion criteria applied were heterogeneous, but generally based on eGFR (n=442, 50%) or serum creatinine (n=324, 37%) (Figure 1c). The exclusion of patients with CKD for individual drug groups is illustrated in Figure 1b.

**Figure 1:**
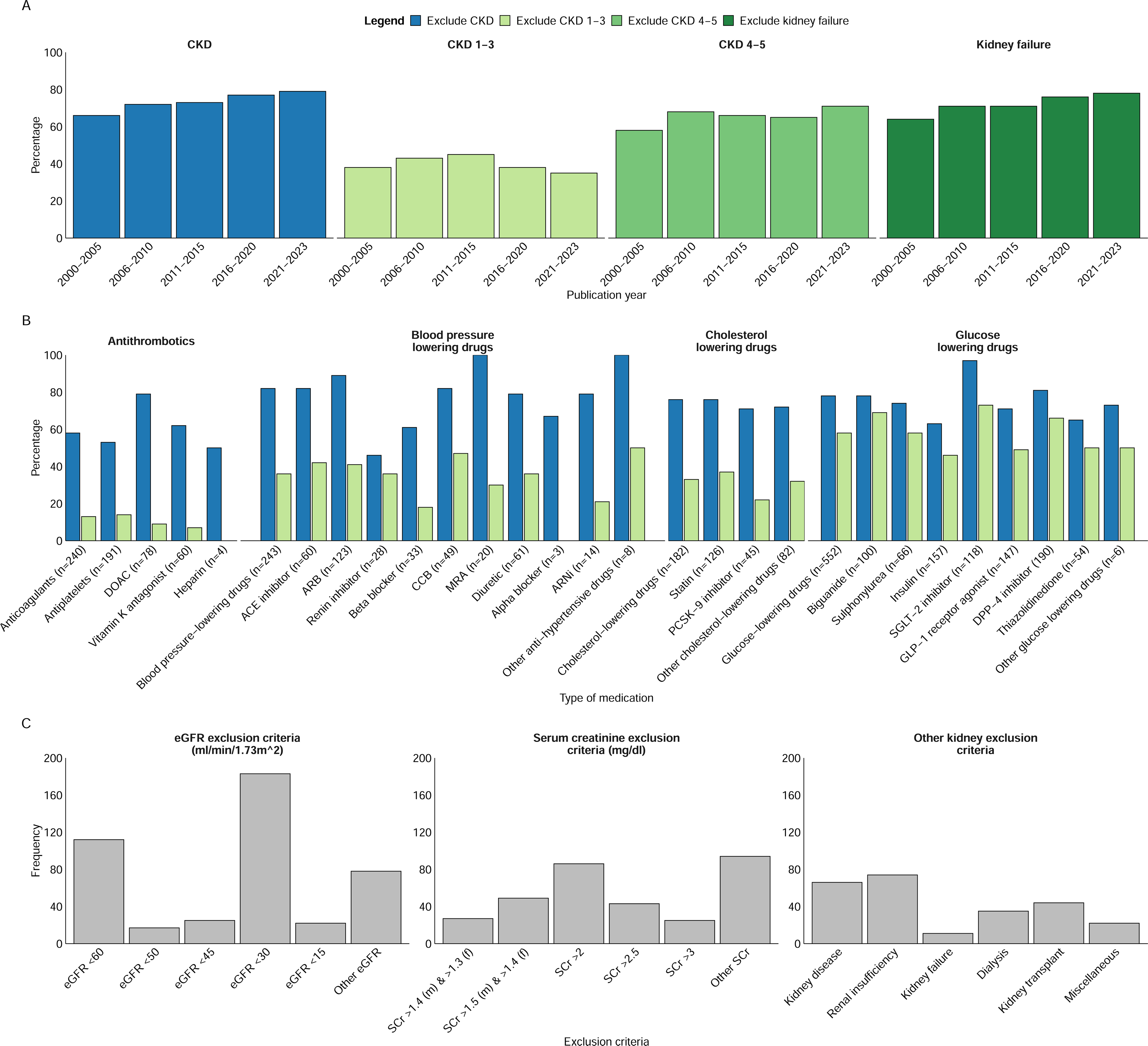
Overview of the exclusion of people with CKD from cardiovascular RCTs. Panel A shows the percentage of RCTs that exclude (subgroups of) people with different stages of CKD over time; panel B shows the exclusion of people with CKD for different types of CVRM medications; panel C shows the most commonly used exclusion criteria for eGFR (ml/min/1.73m^2^), serum creatinine (mg/dL), and other kidney exclusion criteria, respectively. Abbreviations: ACE inhibitor, angiotensin-converting enzyme inhibitor; ARB, angiotensin receptor blocker; CCB, calcium channel blocker; CKD, chronic kidney disease; DOAC, direct oral anticoagulants; DPP-4 receptor agonist, dipeptidyl-peptidase-4 inhibitors; eGFR, estimated glomerular filtration rate; GLP-1 receptor agonist, glucagon-like peptide-1 receptor agonist; MRA, mineralocorticoid receptor antagonist; PSCK-9 inhibitor, proprotein convertase subtilisin/kexin type 9 inhibitor; SCr, serum creatinine; SGLT-2 inhibitor, sodium-glucose cotransporter-2 inhibitor. Other eGFR exclusion criteria were eGFR <40 (n=8); <25 (n=7); >60 and <30, <35 (both n=5); eGFR >50, >60 (both n=3); eGFR <36, >30, >70 and <20, >60 and <15, >60 and <45, creatinine clearance <60% of the normal age-adjusted value (all n=2); eGFR <10, >45 and <20, >45 and <25, >50 and <30, >60 and <25, >75 and, <25, >80 and <20, >90 and <25, >90 and <30, <70, <90, not satisfying eGFR (all n=1). Other serum creatinine exclusion criteria were serum creatinine >1.3 (M) >1.2 (F) (n=7); serum creatinine >1.5 (M) >1.2 (F) (n=6); serum creatinine >1.5 (M) >1.3 (F) (n=5); serum creatinine >1.4, >2.3, elevated serum creatinine level (all n=4); serum creatinine >4, (not further specified; n=3); serum creatinine >1.5 times ULN (M), > 1.4 times ULN (F); >1.5times ULN, >2 times ULN, >3 times ULN, >1.6, >1.8, >2 (M) >1.8 (F), >3 and <1,5 (M), >3 and <1.3 (F) (all n=2); <1,7 (M) <1.5 (F); serum creatinine <2, >2.5 and <1.2 (M), <1.0 (F), >5, and <1.5, >1,7 (M) >1.5 (F), >1.5 (M) >1.1 (F), >1.6 (M) >1.5 (F), >2 (M) >1.5 (F), >2 (M) >1.7 (F), >2.2, >2.4, >2.4 (M) > 2.0 (F), >2.7, >3.5, >6 (all n=1). Other kidney exclusion criteria were not receiving dialysis (n=8); abnormal laboratory results on kidney function tests (not further specified; n=4); serum creatinine or creatinine clearance that was a contraindication to metformin according to the country-specific label (n=2); expected Kidney transplant within 1 year and not on dialysis, patients with serious kidney damage, renal artery stenosis in a solitary kidney or kidney transplant or, one functioning kidney, no kidney transplant, polycystic kidney disease, unsuccessful kidney transplantation (all n=1).

**Figure 2:**
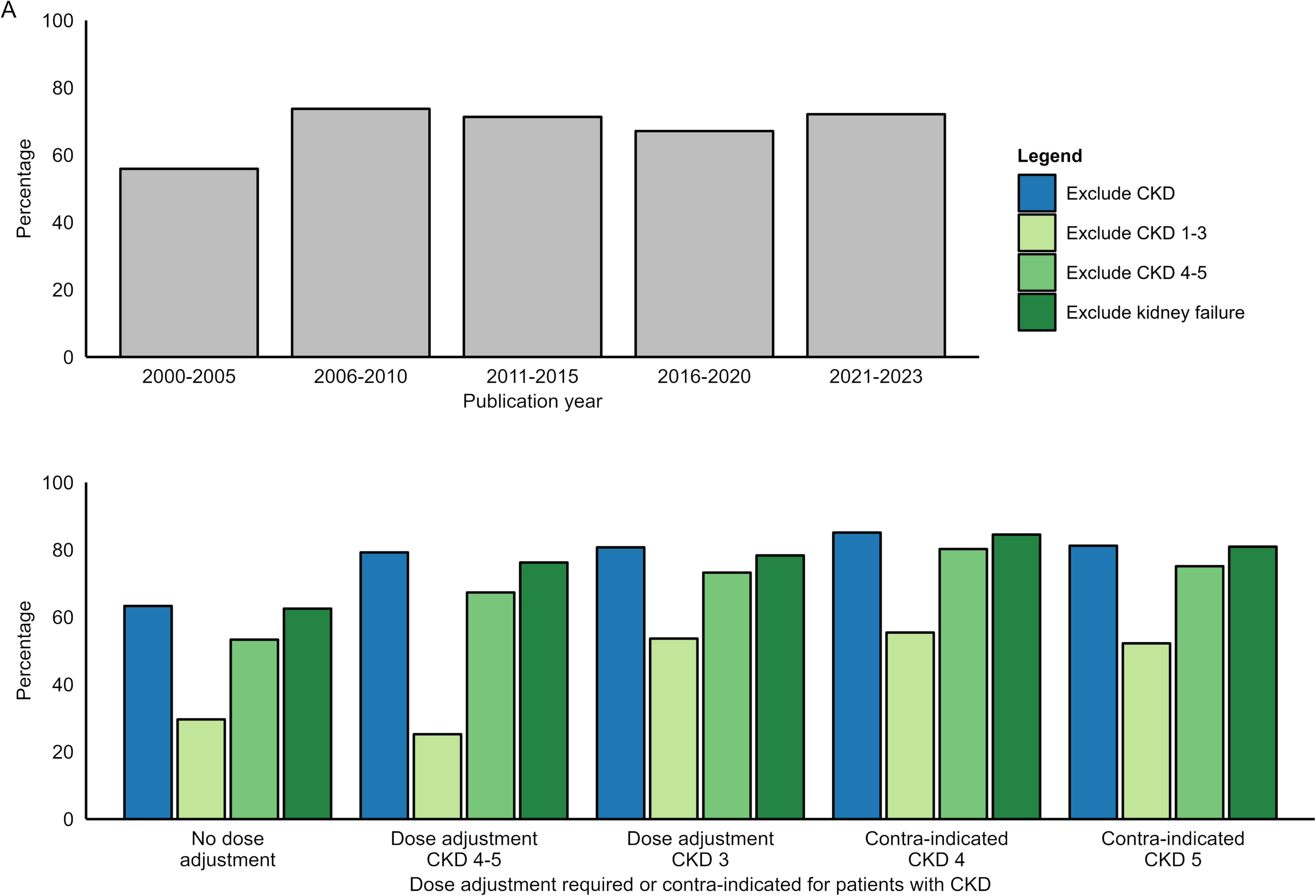
Exclusion of patients with CKD stratified by prescription recommendations for patients with CKD. Panel A shows the percentage of RCTs that exclude different subgroups of patients with CKD stratified by recommendations for dose-adjustment or contra-indications; panel B shows the percentage of RCTs over time where dose adjustment is recommended. Abbreviations: CKD, chronic kidney disease

In 73% of RCTs, more patients with CKD were excluded than expected on safety grounds. Patients with CKD were excluded in 63% of RCTs where no dose adjustment for the interventions on kidney function was required (306/488). The rate of exclusion of patients with CKD was over 80% in RCTs were the dose-adjustments based on kidney function were necessary or interventions were contra-indicated based on kidney function. However, 79% of these RCTs (558/706 RCTs) also excluded more patients with CKD than necessary on safety grounds (Figure 2a).

### Evidence (gaps) for CVRM medications in patients with CKD

In total 158 RCTs (13%) reported results for patients with CKD. Of these RCTs, 34 (3%) only included patients with CKD (4 cholesterol-lowering drugs, 13 blood-pressure lowering drugs, 15 glucose-lowering drugs, and 2 antithrombotic drugs). The percentage of RCTs that reported results for patients with CKD has not improved in the past 20 years (Figure 3b) Analyses for patients with CKD were predominantly performed for composite cardiovascular endpoints (112 RCTs, 66%) in heterogeneous strata (Figure 3a, 4, and 5). Few RCTs conducted analyses for individual cardiovascular endpoints, particularly for heart failure, peripheral arterial disease, and kidney failure (Figure 3a, Supplemental tables 3-9). RCTs that conducted subgroup analyses for patients with CKD had a mean baseline eGFR of 71±12 ml/min/1.73m2, although this parameter was reported in only 40% of the RCTs.

**Figure 3:**
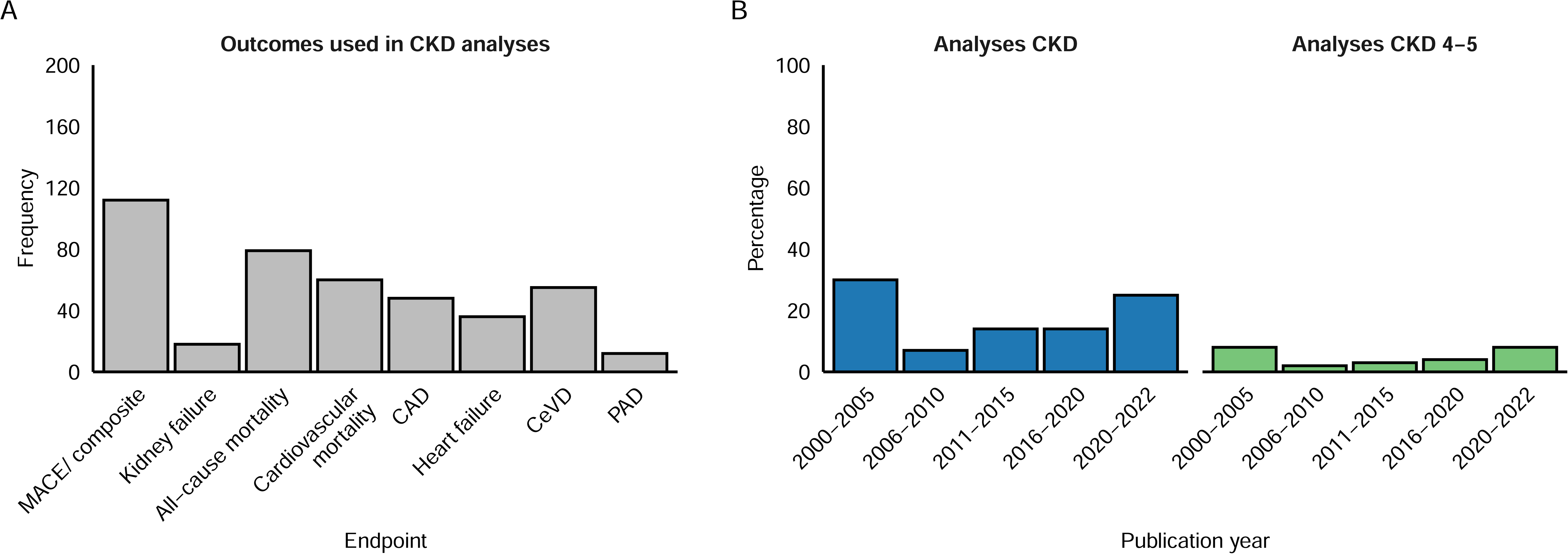
Number of RCTs with analyses for people with CKD for different types of CVRM medications. panel A shows the number of analyses for people with CKD for different cardiovascular endpoints; Panel B shows the percentage of RCTs with analyses for people with any CKD and CKD stage 4-5. Abbreviatons: CAD, coronary artery disease; CeVD, cerebrovascular disease; CKD, chronic kidney disease; MACE, major adverse cardiovascular event; PAD, peripheral arterial disease.

We identified significant gaps in evidence for CVRM medications for all patients with CKD. Evidence gaps were most notable for patients with CKD stage 4-5 for whom 39 RCTs reported results (23 for eGFR <30 ml/min/1.73m^2^, 10895 participants; 15 for dialysis 9516 participants, 1 for kidney transplant recipients; 2102 participants. Figures 4 and 5). An overview of analyses for endpoints other than major adverse cardiovascular events (MACE) is added in the supplementary materials (Supplementary Figures 3-9).

**Figure 4:**
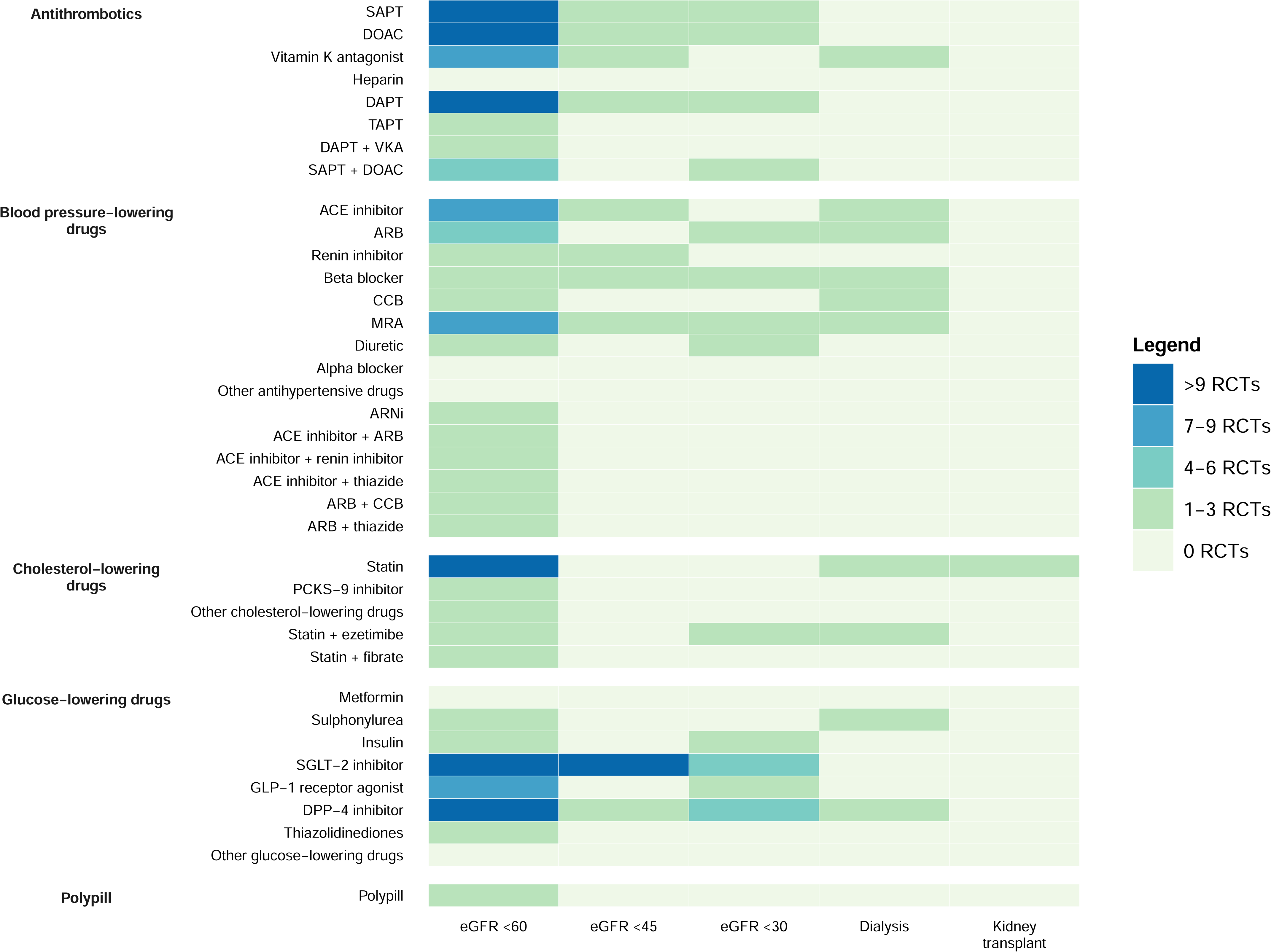
Heat map of (subgroup) analyses for major adverse cardiovascular events for people with different stages of CKD. Abbreviations: ACE-inhibitor, angiotensin-converting enzyme inhibitor; ARB, angiotensin receptor blocker; ARNi, angiotensin receptor-neprilysin inhibitor; CCB, calcium channel blocker; DAPT, double antiplatelet therapy; DOAC, direct oral anticoagulant; DPP-4 inhibitor, dipeptidyl-peptidase-4 inhibitors; eGFR, estimated glomerular filtration rate; GLP-1 receptor agonist, glucagon-like peptide-1 receptor agonist; MRA, mineralocorticoid receptor antagonist; PCSK-9 inhibitor, proprotein convertase subtilisin/kexin type 9 inhibitor; SAPT, single antiplatelet therapy; SGLT-2 inhibitor, sodium-glucose cotransporter-2 inhibitor; TAPT, triple antiplatelet therapy; VKA, vitamin K antagonist.

**Figure 5:**
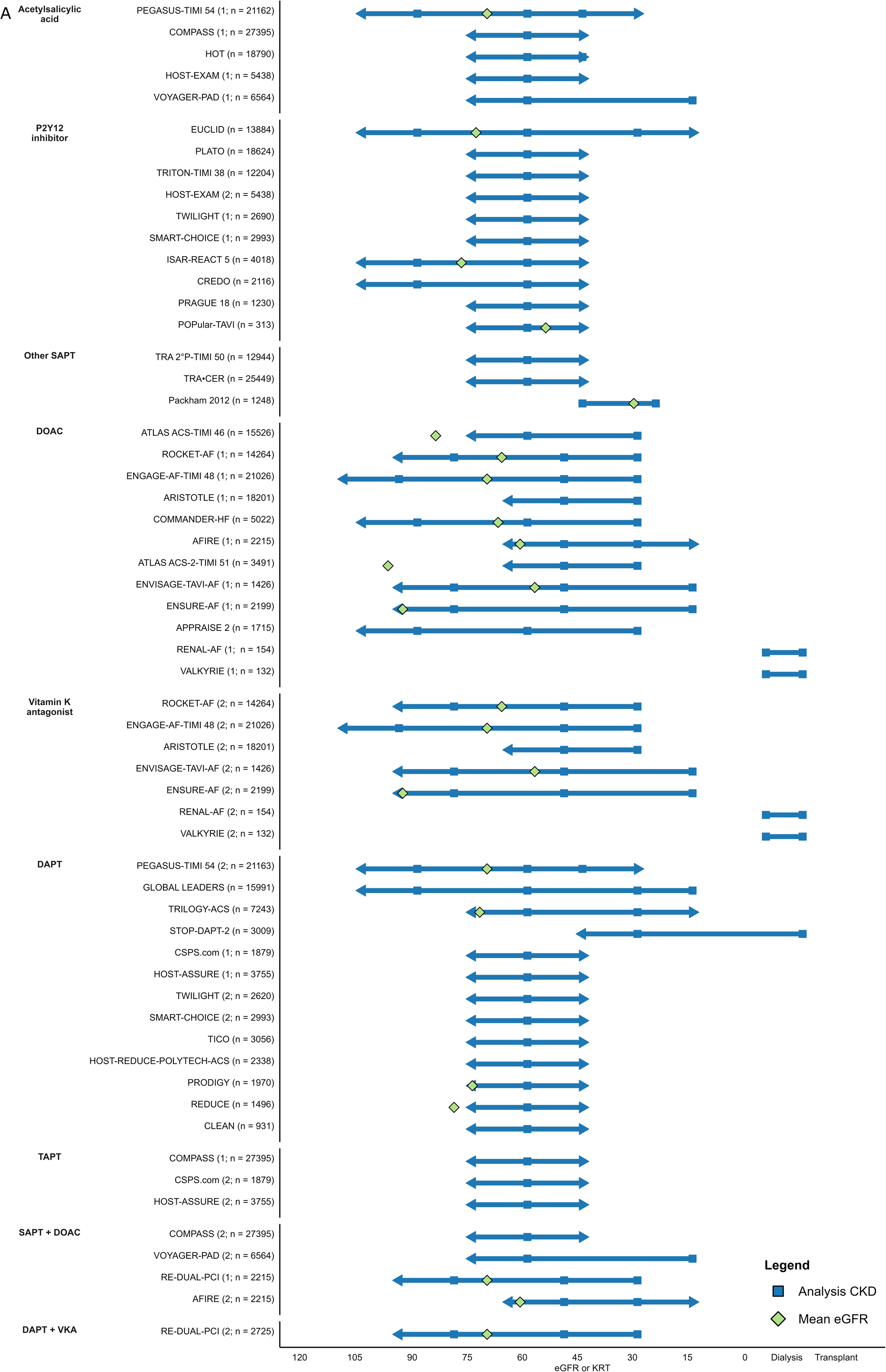
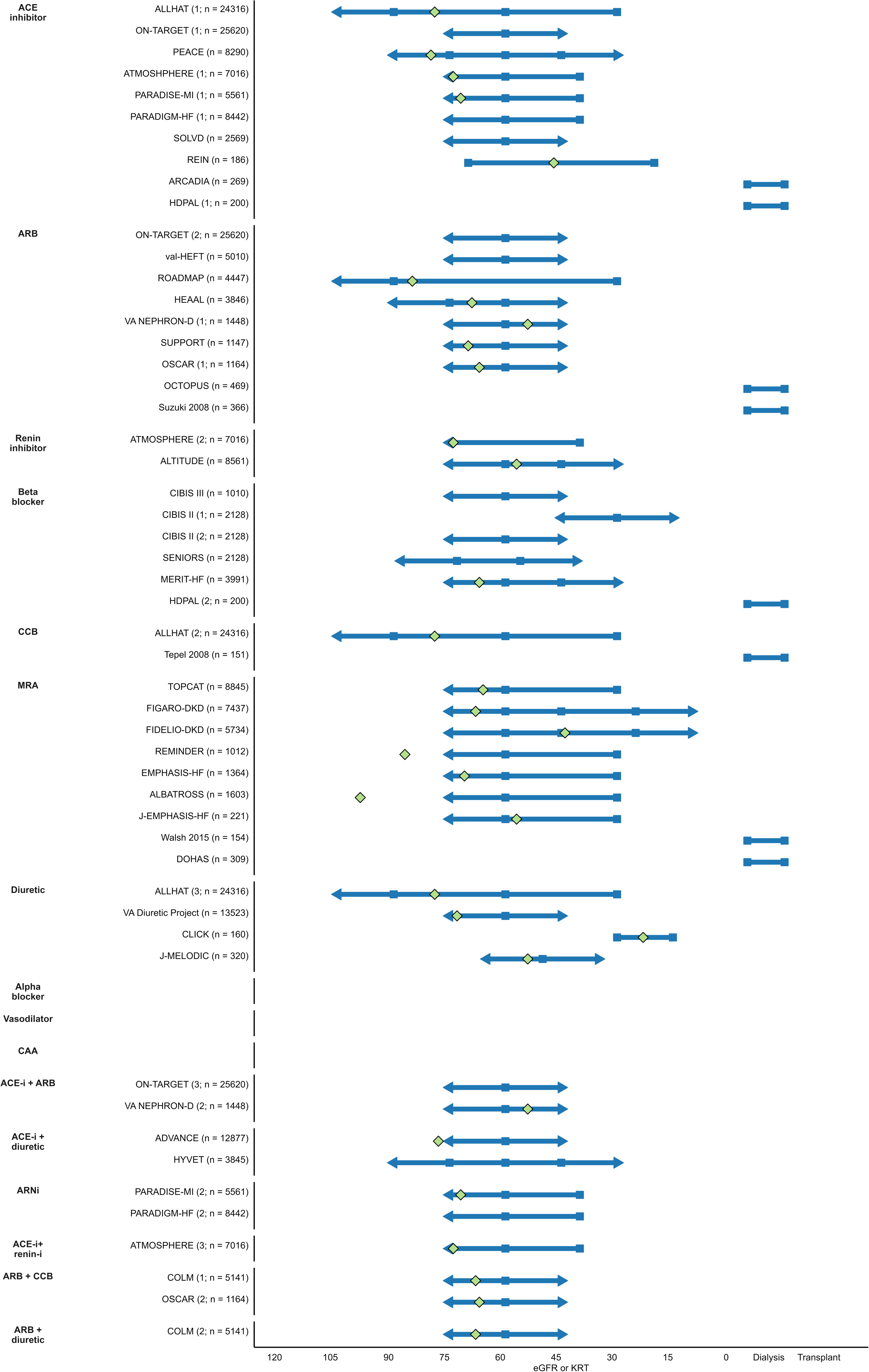
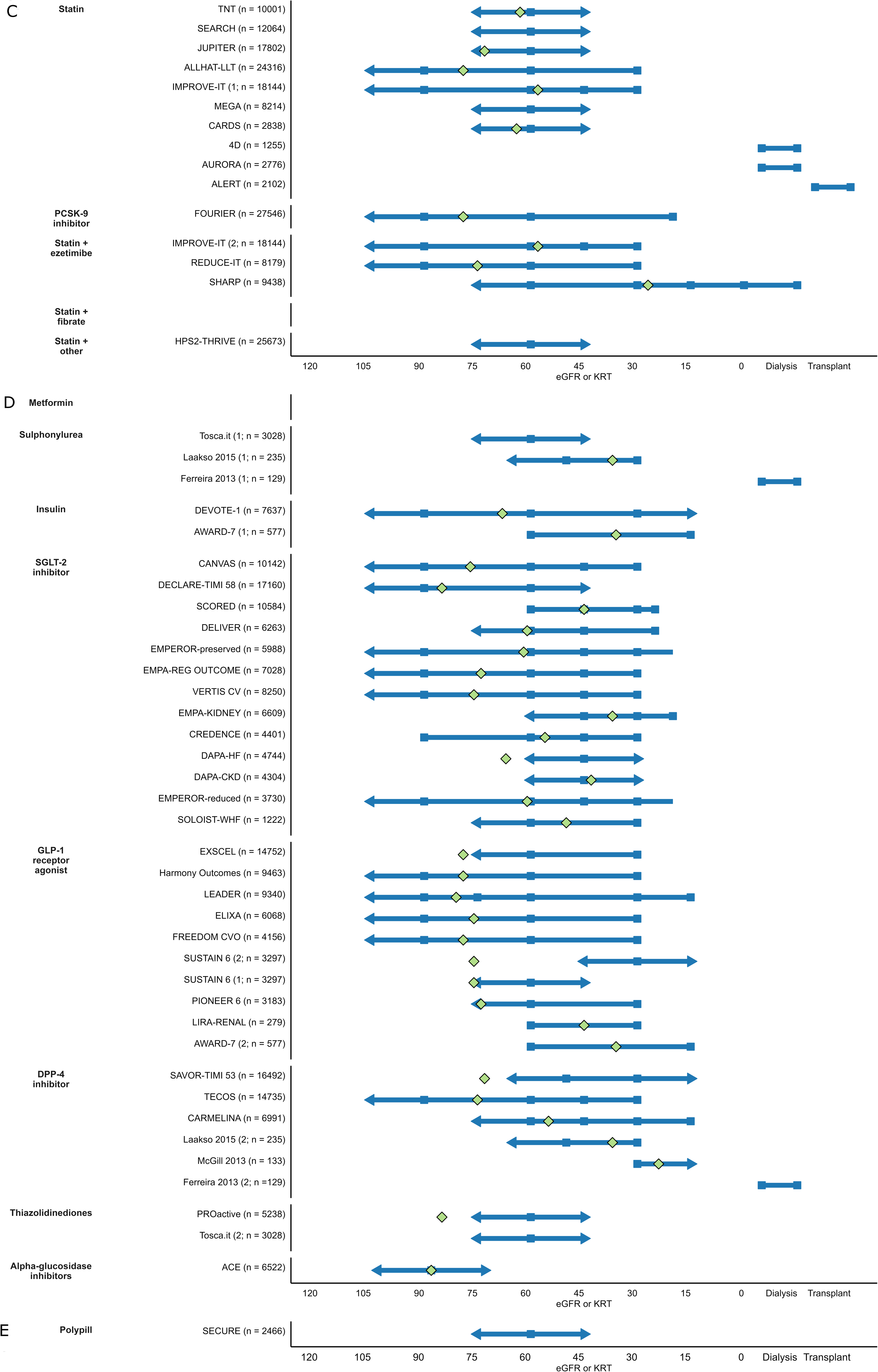
Overview of individual studies with analysis on eGFR or kidney replacement therapy for major adverse cardiovascular events in patients with CKD. Panel A illustrates analyses for antithrombotics; panel B for blood- pressure-lowering drugs; panel C for cholesterol-lowering drugs; panel D for glucose-lowering drugs; and panel E for polypills. Abbreviations: ACE-inhibitor/ACE-I, angiotensin-converting enzyme; ARB, angiotensin receptor blocker; CAA, centrally-acting antihypertensive; CCB, calcium channel blocker; CKD, chronic kidney disease; DAPT, double antiplatelet therapy; DOAC, direct oral anticoagulants; DPP-4 inhibitor, dipeptidyl-peptidase-4 inhibitor; eGFR, estimated glomerular filtration rate; GLP-1 receptor agonist, glucagon-like peptide-1 receptor agonist; KRT, kidney replacement therapy; MRA, mineralocorticoid receptor antagonist; PCSK-9 inhibitor, proprotein convertase subtilisin/kexin type 9 inhibitor; SAPT, single antiplatelet therapy; SGLT-2 inhibitor, sodium-glucose cotransporter-2 inhibitor; TAPT, triple antiplatelet therapy; VKA, vitamin K antagonist.

#### Blood pressure-lowering drugs

Most RCTs evaluating blood pressure-lowering drugs for the prevention of major adverse cardiovascular events (MACE) for patients with CKD studied angiotensin-converting enzyme-inhibitors (ace-inhibitors, angiotensin receptor blockers (ARBs) and mineralocorticoid receptor blockers (MRAs) (Figures 4 and 5). For patients with an eGFR <60 ml/min/1.73m^2^, each antihypertensive was evaluated in at least 1 RCT except for alpha-blockers. ARBs, beta blockers and MRAs were evaluated for both patients with an eGFR <30 ml/min/1.73m^2^ and patients receiving dialysis. Ace-inhibitors and calcium channel blockers were also evaluated for patients receiving dialysis and thiazides for patients with an eGFR <30 ml/min/1.73m^2^. Other antihypertensives were not evaluated in these populations. None of the RCTs evaluated the efficacy of antihypertensives for patients with a kidney transplant (Figures 4 and 5).

#### Cholesterol-lowering drugs

The efficacy of cholesterol-lowering drugs in patients with CKD on MACE was evaluated almost exclusively for statins but barely for other cholesterol-lowering drugs (Figures 3 and 4). Statins were evaluated for patients with an eGFR <60 ml/min/1.73m^2^ as monotherapy or in combination with ezetimibe. PCSK-9 (proprotein convertase subtilisin/kexin type 9) inhibitors, niacin, and icosapent ethyl were also evaluated for this population but not for patients with CKD stage 4-5. For patients with an eGFR <30 ml/min/1.73m^2^, the combination of statins with ezetimibe was the only therapy evaluated. For patients receiving dialysis, statins were evaluated as monotherapy and combination with ezetimibe. For patients with a kidney transplant, statin monotherapy was evaluated but none of the other drugs (Figures 4 and 5).

#### Antiplatelets and anticoagulants

Of all CVRM medications, antiplatelets and anticoagulants were studied most frequently for patients with CKD (Figures 3 and 4). At least 10 RCTs evaluated the efficacy of single antiplatelet therapy, double antiplatelet therapy (SAPT and DAPT, respectively), and DOACs for patients with CKD. However, these analyses were largely restricted to patients with an eGFR <60 ml/min/1.73m^2^ (Figures 3 and 4). The efficacy of triple antiplatelet therapy or combinations of DAPT or SAPT with vitamin K antagonist (VKAs) or DOACs was evaluated in a handful of RCTs and only one of these RCTs evaluated the efficacy of these interventions in patients with an eGFR <60 ml/min/1.73m^2^. For patients with an eGFR <30 ml/min/1.73m^2^, the efficacy of SAPT, DAPT, DOACs, and a combination of DOACs with SAPT was evaluated. For patients receiving dialysis, evidence was limited to the comparison of DOACs with VKAs and the efficacy of antiplatelets was not evaluated at all. None of the RCTs evaluated the efficacy of antiplatelets and anticoagulants for patients with a kidney transplant (Figures 4 and 5).

#### Glucose-lowering drugs

The efficacy of newer glucose-lowering drugs (SGLT-1 inhibitors and GLP-1 receptor agonists) on MACE for patients with CKD was evaluated in multiple RCTs. However, hardly any evidence was available for older glucose-lowering drugs (metformin, sulphonylureas, and insulin; Figures 3 and 4). Similarly to antiplatelets and anticoagulants, evidence was mainly restricted to patients with an eGFR <60 ml/min/1.73m^2^. For patients with an eGFR <30 ml/min/1.73m^2^, the efficacy of SGLT-2 inhibitors, DPP-4 inhibitors, GLP-1 receptor agonists, and insulin was evaluated. For patients receiving dialysis, the efficacy DPP-4 inhibitors was compared with sulphonylureas but no other drugs were studied. None of the RCTs evaluated the efficacy of glucose-lowering drugs in patients with a kidney transplant (Figures 4 and 5).

## Discussion

In our systematic review, including almost 1200 RCTs and over 2.2 million patients, we have identified no improvement in the underrepresentation of patients with CKD over the past two decades. On the contrary, since 2000 the number of cardiovascular RCTs that excluded subgroups of patients with CKD has increased. Exclusion criteria were heterogenous and cardiovascular RCTs consistently excluded a larger number of patients with CKD than would be anticipated on safety grounds. In addition, only 13% of included cardiovascular RCTs evaluated the efficacy of CVRM medications for patients with CKD, mostly in subgroup analyses, resulting in evidence gaps across all CVRM medications and subgroups of patients with CKD.

The persistently high proportion of RCTs that exclude patients with CKD in the past 20 years cannot be attributed to a rise in the number of RCTs requiring dose-adjustment on kidney function for CVRM medications as the proportion of RCTs requiring such adjustment remained stable. While excluding patients with CKD from such RCTs due to safety concerns can be justifiable, the significantly more stringent kidney exclusion criteria compared to prescription thresholds in clinical practice suggest there are additional reasons for excluding patients with CKD. Practical issues like the necessity for dose adjustments, concerns about heterogeneity in treatment effects, or a limited life expectancy could also discourage clinical trialists from including patients with CKD in their RCTs.

The evidence gaps for patients with CKD in cardiovascular RCTs can be traced back to the ongoing widespread exclusion of this population. Excluding patients with CKD due to possible treatment heterogeneity or initial safety concerns does not necessarily lead to evidence gaps provided that separate RCTs are conducted to assess the efficacy of medications for patients with CKD. However, in practice, only 0.25% of included cardiovascular RCTs was conducted specifically for patients with CKD.

### Implications for practice of the evidence gaps for CVRM medications in patients with CKD

Currently, most RCTs which evaluated CVRM medications for patients with CKD focussed on those with CKD stage 3. For patients with CKD stage 4-5, which comprise 10% of the patients with CKD (i.e. 85 million patients) (41), analyses were often absent, particularly for those with a kidney transplant. The lack of RCTs assessing the efficacy of CVRM medications for patients with CKD means that, in practice, practitioners must resort to extrapolating results from RCTs conducted in other populations, assuming that the treatment effects are comparable. However, this assumption is increasingly less likely to hold for patients with more advanced CKD stages where CKD-specific risk factors like vascular calcification, uraemia, chronic inflammation, and immunosuppressive therapy to prevent graft rejection, combined with a very high risk and reduced life expectancy can modify the treatment effect (42, 43).

The complexity of extrapolating results to patients with CKD is illustrated by statins. Although these drugs reduce cardiovascular risk in patients with an eGFR <60 ml/min/1.73m^2^, their efficacy has not been demonstrated in individuals with kidney failure (44). The lack of RCTs conducted in patients between these ends of the CKD spectrum makes it impossible to determine the tipping point where statins lose their benefits. Consequently, patients may unintentionally be over- or undertreated since the balance between benefits and side effects remains unknown.

In addition to an absolute lack of RCTs, limitations in the analyses further hamper CVRM treatment for patients with CKD. Heterogeneity in exclusion criteria and inadequate reporting of baseline kidney function reduce the comparability of RCTs while the small sample size of strata leads to underpowered analyses and imprecision in effect estimates. Furthermore, the lack of RCTs evaluating the effect of CVRM medications on individual cardiovascular endpoints and kidney endpoints in patients with CKD means that the efficacy of these drugs on these individual endpoints remains unknown. More importantly, stratifying RCT cohorts breaks randomisation and can introduce confounding due to clustering of other CVD risk factors in patients with CKD (45, 46). These limitations are likely to amount in a GRADE recommendation of low or very low certainty of evidence for most CVRM medications in patients with CKD, meaning that their efficacy might be markedly different from the estimated effect (47).

### Implications for research

An increasing prevalence of CKD (including dialysis and transplantation), the widespread prescription of CVRM medications to this population, and uncertainty about the efficacy of various CVRM medications in patients with CKD underscore the urgency of adequate representation of patients with CKD in cardiovascular RCTs (48, 49). Despite efforts of the United States Food and Drug Administration (FDA) and European Medicine Agency (EMA) to promote the enrolment of patients with CKD and numerous reviews and editorials addressing this issue, the representation of patients with CKD in cardiovascular RCTs has not improved in the past twenty years (12, 13, 50, 51). Bridging the evidence gap for treatment of cardiovascular risk in patients with CKD is the responsibility of different stakeholders including pharmaceutical companies, medicine regulatory authorities, scientific societies, and funding bodies and starts with the proportional inclusion of patients with CKD to enable (subgroup) analyses on kidney disease. New evidence for patients with CKD stage 4-5 (including those receiving kidney replacement therapy) should be prioritized considering the evidence gaps are largest for this population. However, such analyses are only feasible if the enrolment of these patient improves and separate RCTs are conducted for them. Additionally, more evidence is needed on the efficacy of CVRM medications on individual cardiovascular and kidney endpoints. Innovative RCT designs like adaptive platform trials based on a master protocol might be an attractive means to rapidly generate evidence for a range of treatment strategies for different groups of patients with CKD. Emulated target trials with real-world data present another opportunity to fill evidence gaps for the efficacy of CVRM medications in patients with CKD, especially for drugs regularly prescribed in practice where conducting new RCTs is prohibitively expensive and time consuming (52).

### Strengths and limitations

The main strength of this review is its methodological rigour: we performed screening and data extraction in pairs and published our protocol a priori (17). Unlike previous reviews, we included all CVRM medications recommended in the major American and European guidelines and RCTs evaluating these medications both in primary and secondary prevention settings (18–39). Furthermore, this is the first study to compare the exclusion of patients with CKD with the recommended thresholds for dose-adjustment or contra-indications on kidney function. Limitations of this systematic review are that we might have underestimated the exclusion of patients with CKD in RCTs with ambiguous exclusion criteria such as chronic disease or life-limiting disease and have missed RCTs not registered in ClinicalTrials.gov. However, we are confident the number of missed RCTs is small since the sensitivity of searches in trial registries and electronic databases has demonstrated to be comparable (53). Moreover, our validation study showed that our search strategy identified almost all eligible RCTs from a bibliographic database search. Searching for RCTs registered in or after 2000 may appear a limitation as we did not include most RCTs published before this date without retrospective registration. However, these older RCTs are less relevant for contemporary clinical practice and guidelines recommendations as more comprehensive CVRM care and new therapies have vastly improved patients’ prognosis (54).

### Conclusion

The representation of patients with CKD in cardiovascular RCTs has not improved in the past twenty years. Cardiovascular RCTs systematically excluded more patients with CKD than expected on safety grounds. The lack of cardiovascular RCTs that report results for patients with CKD has resulted in significant evidence gaps for the efficacy of most CVRM medications in patients with CKD, particularly for those with CKD stage 4-5.

## Other information

Registration and protocol

PROSPERO: CRD42022296746

Protocol: Colombijn JMT, Idema DL, van der Braak K, Spijker R, Meijvis SCA, Bots ML, et al. Evidence for pharmacological interventions to reduce cardiovascular risk for patients with chronic kidney disease: a study protocol of an evidence map. Syst Rev. 2022;11(1):238.

## Availability of data, code, and other materials

All data in this review are based on publicly available articles. Dataset and syntax are available upon request

## Supplementary Figures

**Supplementary Figure 1:** Flow chart of literature search.

**Supplementary Figure 2:** Exclusion of patients with CKD stratified by prescription recommendations for patients with CKD.

**Supplementary Figure 3:** Heat map of (subgroup) analyses for all-cause mortality for people with different stages of CKD.

Abbreviations: ACE-inhibitor, angiotensin-converting enzyme inhibitor; ARB, angiotensin receptor blocker; ARNi, angiotensin receptor-neprilysin inhibitor; CCB, calcium channel blocker; DAPT, double antiplatelet therapy; DOAC, direct oral anticoagulant; DPP-4 inhibitor, dipeptidyl-peptidase-4 inhibitors; eGFR, estimated glomerular filtration rate; GLP-1 receptor agonist, glucagon-like peptide-1 receptor agonist; MRA, mineralocorticoid receptor antagonist; PCSK-9 inhibitor, Proprotein convertase subtilisin/kexin type 9 inhibitor; SAPT, single antiplatelet therapy; SGLT-2 inhibitor, sodium-glucose cotransporter-2 inhibitor; TAPT, triple antiplatelet therapy; VKA, vitamin K antagonist.

**Supplementary Figure 4:** Heat map of (subgroup) analyses for cardiovascular mortality for people with different stages of CKD.

Abbreviations: ACE-inhibitor, angiotensin-converting enzyme inhibitor; ARB, angiotensin receptor blocker; ARNi, angiotensin receptor-neprilysin inhibitor; CCB, calcium channel blocker; DAPT, double antiplatelet therapy; DOAC, direct oral anticoagulant; DPP-4 inhibitor, dipeptidyl-peptidase-4 inhibitors; eGFR, estimated glomerular filtration rate; GLP-1 receptor agonist, glucagon-like peptide-1 receptor agonist; MRA, mineralocorticoid receptor antagonist; PCSK-9 inhibitor, Proprotein convertase subtilisin/kexin type 9 inhibitor; SAPT, single antiplatelet therapy; SGLT-2 inhibitor, sodium-glucose cotransporter-2 inhibitor; TAPT, triple antiplatelet therapy; VKA, vitamin K antagonist.

**Supplementary Figure 5:** Heat map of (subgroup) analyses for coronary artery disease for people with different stages of CKD.

Abbreviations: ACE-inhibitor, angiotensin-converting enzyme inhibitor; ARB, angiotensin receptor blocker; ARNi, angiotensin receptor-neprilysin inhibitor; CCB, calcium channel blocker; DAPT, double antiplatelet therapy; DOAC, direct oral anticoagulant; DPP-4 inhibitor, dipeptidyl-peptidase-4 inhibitors; eGFR, estimated glomerular filtration rate; GLP-1 receptor agonist, glucagon-like peptide-1 receptor agonist; MRA, mineralocorticoid receptor antagonist; PCSK-9 inhibitor, Proprotein convertase subtilisin/kexin type 9 inhibitor; SAPT, single antiplatelet therapy; SGLT-2 inhibitor, sodium-glucose cotransporter-2 inhibitor; TAPT, triple antiplatelet therapy; VKA, vitamin K antagonist.

**Supplementary Figure 6:** Heat map of (subgroup) analyses for heart failure for people with different stages of CKD.

Abbreviations: ACE-inhibitor, angiotensin-converting enzyme inhibitor; ARB, angiotensin receptor blocker; ARNi, angiotensin receptor-neprilysin inhibitor; CCB, calcium channel blocker; DAPT, double antiplatelet therapy; DOAC, direct oral anticoagulant; DPP-4 inhibitor, dipeptidyl-peptidase-4 inhibitors; eGFR, estimated glomerular filtration rate; GLP-1 receptor agonist, glucagon-like peptide-1 receptor agonist; MRA, mineralocorticoid receptor antagonist; PCSK-9 inhibitor, Proprotein convertase subtilisin/kexin type 9 inhibitor; SAPT, single antiplatelet therapy; SGLT-2 inhibitor, sodium-glucose cotransporter-2 inhibitor; TAPT, triple antiplatelet therapy; VKA, vitamin K antagonist.

**Supplementary Figure 7:** Heat map of (subgroup) analyses for cerebrovascular disease for people with different stages of CKD.

Abbreviations: ACE-inhibitor, angiotensin-converting enzyme inhibitor; ARB, angiotensin receptor blocker; ARNi, angiotensin receptor-neprilysin inhibitor; CCB, calcium channel blocker; DAPT, double antiplatelet therapy; DOAC, direct oral anticoagulant; DPP-4 inhibitor, dipeptidyl-peptidase-4 inhibitors; eGFR, estimated glomerular filtration rate; GLP-1 receptor agonist, glucagon-like peptide-1 receptor agonist; MRA, mineralocorticoid receptor antagonist; PCSK-9 inhibitor, Proprotein convertase subtilisin/kexin type 9 inhibitor; SAPT, single antiplatelet therapy; SGLT-2 inhibitor, sodium-glucose cotransporter-2 inhibitor; TAPT, triple antiplatelet therapy; VKA, vitamin K antagonist.

**Supplementary Figure 8:** Heat map of (subgroup) analyses for peripheral arterial disease for people with different stages of CKD.

Abbreviations: ACE-inhibitor, angiotensin-converting enzyme inhibitor; ARB, angiotensin receptor blocker; ARNi, angiotensin receptor-neprilysin inhibitor; CCB, calcium channel blocker; DAPT, double antiplatelet therapy; DOAC, direct oral anticoagulant; DPP-4 inhibitor, dipeptidyl-peptidase-4 inhibitors; eGFR, estimated glomerular filtration rate; GLP-1 receptor agonist, glucagon-like peptide-1 receptor agonist; MRA, mineralocorticoid receptor antagonist; PCSK-9 inhibitor, Proprotein convertase subtilisin/kexin type 9 inhibitor; SAPT, single antiplatelet therapy; SGLT-2 inhibitor, sodium-glucose cotransporter-2 inhibitor; TAPT, triple antiplatelet therapy; VKA, vitamin K antagonist.

**Supplementary Figure 9:** Heat map of (subgroup) analyses kidney failure for people with different stages of CKD.

Abbreviations: ACE-inhibitor, angiotensin-converting enzyme inhibitor; ARB, angiotensin receptor blocker; ARNi, angiotensin receptor-neprilysin inhibitor; CCB, calcium channel blocker; DAPT, double antiplatelet therapy; DOAC, direct oral anticoagulant; DPP-4 inhibitor, dipeptidyl-peptidase-4 inhibitors; eGFR, estimated glomerular filtration rate; GLP-1 receptor agonist, glucagon-like peptide-1 receptor agonist; MRA, mineralocorticoid receptor antagonist; PCSK-9 inhibitor, Proprotein convertase subtilisin/kexin type 9 inhibitor; SAPT, single antiplatelet therapy; SGLT-2 inhibitor, sodium-glucose cotransporter-2 inhibitor; TAPT, triple antiplatelet therapy; VKA, vitamin K antagonist.

